# The quality–failure paradox in extracellular vesicle therapeutics: quantitative benchmarking of 152 clinical trials against CAR-T cell therapies

**DOI:** 10.64898/2026.07.20.26358510

**Authors:** Konstantin Glebov, Natasa Zarovni

## Abstract

The stall in clinical translation of extracellular vesicle (EV) therapeutics requires addressing critical and hidden causes of attrition and delays. Despite more than 150 registered clinical trials and fifteen years of clinical investigation, small EV (sEV) therapeutics have yielded zero FDA approvals—a translational deficit that has received remarkably little quantitative scrutiny. We systematically evaluated 783 EV-related clinical trials (152 of which therapeutic) registered through December 2025, benchmarking the sEV pipeline against 1,131 CAR-T cell therapy trials (ex-China) that share the “process is the product” manufacturing constraint, yet have delivered six FDA-approved products. A quality–failure paradox emerges, in which industry-sponsored sEV trials exhibit 39% higher composite methodological quality (quality index 2.49 vs. 1.79) yet fail at approximately five-fold the rate of academic programmes (28.6% vs. 5.1%; OR=7.40, 95% CI 2.5–22.3, p=0.0004). Cross-modality replication in CAR-T trials confirms the direction of this association (OR=2.51, p<0.001; Cochran– Mantel–Haenszel pooled OR=2.76, p<0.001). Registry abandonment—affecting 24% of academic sEV trials—constitutes a hidden failure mode that, when reclassified, dissolves the apparent paradox. Premature clinical entry of incompletely defined products, rather than insufficient methodological rigour, represents the central constraint on sEV translational progress. The opacity of research data, inadequate pre clinical evaluation, and failure to report negative (null) findings in clinical trials create a hidden crisis that drains hundreds of millions invested by public and private sector—eroding the return on investment—and traps progress in cycles of duplicated effort, wasteful resource allocation, and missed learning opportunities.

## INTRODUCTION

Extracellular vesicles—micro- and nanoscale membrane-enclosed particles released by virtually all cell types—have attracted sustained attention as a next-generation therapeutic platform, with preclinical data demonstrating tissue repair, immunomodulation, and drug delivery capabilities that would, if translatable, address unmet needs across oncology, neurology, and regenerative medicine (1, 2). The convergence of favourable biological properties (low immunogenicity, capacity to cross biological barriers, natural cargo-loading mechanisms), with claimed scalable biomanufacturing potential has positioned small extracellular vesicles (sEVs) as candidates for both direct therapeutic application, and as drug delivery vehicles (3, 4). The field’s promise, however, stands in stark contrast to its regulatory output: as of December 2025, not a single sEV-based drug products (DPs) has received marketing authorisation from the FDA, EMA, or any major regulatory agency, a deficit that persists despite the registration of more than 150 therapeutic clinical trials spanning a decade of scientific investigation and the investment of substantial public and private capital.

Multiple prior systematic reviews have catalogued the expanding sEV trial landscape—documenting growth in registration volume, geographic diversity, and indication breadth—but have largely abstained from evaluating translational readiness or diagnosing why volumetric expansion has not produced regulatory milestones (5-7). The implicit assumption in much of this literature is that the sEV field remains “early-stage” and that pipeline maturation will follow a predictable trajectory from Phase 1 dose-finding through registration-quality trials. Our data challenge this assumption directly: fourteen-fold growth in annual trial registrations between 2018 and 2024 has occurred without commensurate advancement in phase distribution, with 40% of all therapeutic trials remaining at Phase 1 and only 2.0% reaching Phase 3. The absence of pipeline maturation despite volumetric growth suggests that structural constraints—rather than insufficient investment or regulatory intransigence—impede translational progress, and that identifying these constraints requires analytical frameworks distinct from those employed in prior descriptive surveys.

That the regulatory pathway itself is navigable—rather than structurally prohibitive—is demonstrated by several programmes that have achieved IND clearance and progressed through dose-escalation, including Codiak BioSciences (engineered exosomes displaying STING agonist for solid tumours), Exo Biologics (allogeneic MSC-EVs for inflammatory conditions), Aruna Bio (neural-derived EVs for neurological indications), Aegle Therapeutics (burn wound EVs that received Regenerative Medicine Advanced Therapy designation from FDA), and ILIAS Biologics (optogenetically loaded EVs with engineered surface ligands). These precedents establish that sEV products can satisfy chemistry, manufacturing, and controls (CMC) requirements, preclinical toxicology packages, and safety monitoring frameworks sufficient for human dosing—yet none has completed a pivotal efficacy trial, and several have ceased operations entirely. The translational bottleneck lies not in regulatory gatekeeping *per se,* but in the field’s collective inability to generate definitive efficacy data that appropriately supports marketing applications. The raised question is whether the impediment is fundamentally methodological, product-definitional, or structural.

To contextualise this bottleneck, we selected chimeric antigen receptor T-cell (CAR-T) therapy as a structural comparator—not because the two modalities share pharmacological mechanism, but because they share a manufacturing constraint that the ICH Q5 guideline series articulates as the “process is the product” principle (8–12). In both CAR-T and sEV therapeutics, product identity, potency, and safety are inseparable from the specific manufacturing process used to generate them; alterations of cell source, culture conditions, isolation methodology, or formulation can yield a functionally distinct product even when the starting material is nominally identical. CAR-T therapy resolved this constraint through product-specific clinical development—tisagenlecleucel (13), axicabtagene ciloleucel (14), lisocabtagene maraleucel, and their successors each represent a defined product with a locked manufacturing process, enabling the consistent lot-to-lot characterisation required by regulatory frameworks. The sEV field has largely not achieved this resolution: approximately 80% of therapeutic trials employ unmodified mesenchymal stromal cell-derived EVs with pleiotropic, incompletely characterised mechanisms of action, while manufacturing processes vary substantially across sites without formal comparability assessments and lock of distinctive protocols.

We therefore pursued three hypotheses that, taken together, aim to diagnose why volumetric clinical expansion has not evolved into regulatory and commercial milestones. First, that the quality–failure paradox—whereby industry trials exhibit higher methodological rigour yet higher failure rates—reflects the functioning of a quality infrastructure as a detection mechanism rather than a preventive one, such that well-designed trials identify futility in products that were insufficiently defined prior to clinical entry. Second, that this association replicates across other therapy modalities sharing the process-is-the-product constraint, suggesting a structural phenomenon inherent to complex biotherapeutic development rather than a deficiency specific to sEV biology or manufacturing. Third, that the true rate of translational non-productivity in the sEV field substantially exceeds the observed failure rate when registry abandonment and status ambiguity are accounted for, and that this hidden attrition concentrates disproportionately in academic programmes—implying that the field’s translational inefficiency is distributed more broadly than headline failure rates would suggest.

## RESULTS

### Trial landscape and phase distribution

Search of ClinicalTrials.gov identified 783 clinical trials involving extracellular vesicles registered through December 2025, of which 152 (19.4%) met criteria for therapeutic intent—defined as interventional administration of EVs or EV-based products with a primary efficacy or safety endpoint— while 548 (70.0%) were diagnostic or biomarker studies and 83 (10.6%) addressed other applications including cosmetic, nutritional supplement, or device-related uses (Figure 1A). The therapeutic portfolio spans oncology, regenerative medicine, neurology, autoimmunity, respiratory disease (dominated by COVID 19 ARDS), wound healing, and dermatology — a breadth that reflects the pleiotropic mechanisms of mesenchymal stromal cell–derived sEVs but also reveals opportunistic indication picking across the field’s clinical development strategy. Among therapeutic trials, academic institutions sponsored 107 (70.4%), industry 35 (23.0%), and government agencies 10 (6.6%), representing a 2.3-fold enrichment of industry participation relative to the overall sEV trial population where industry accounts for only 10.2% of registrations (Figure 1B).

**Figure 1.**
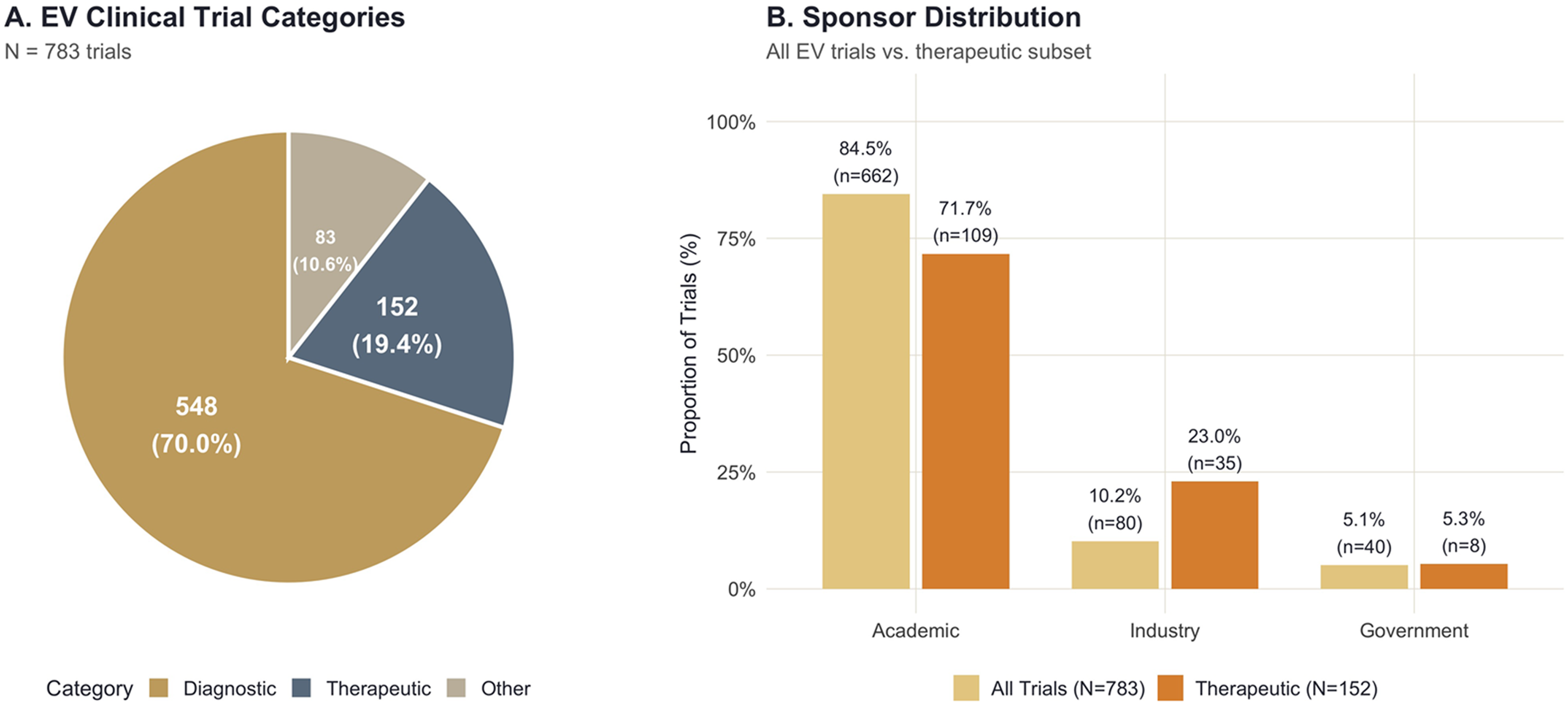
Extracellular Vesicle Clinical Trial Landscape. (A) Distribution of 783 EV-related clinical trials by primary category (diagnostic, therapeutic, other). (B) Sponsor distribution comparing all EV trials with the therapeutic subset, demonstrating 2.3-fold enrichment of industry participation in therapeutic applications.

Phase distribution within the therapeutic subset reveals persistent early-stage concentration without evidence of temporal maturation (Figure 2A). Of 152 trials, 61 (40.1%) are designated Phase 1 or Early Phase 1, 36 (23.7%) Phase 1/Phase 2, 17 (11.2%) Phase 2, and only 3 (2.0%) Phase 3—with the remaining 31 (20.4%) lacking any phase designation, a proportion significantly exceeding that observed in CAR-T trials (9.2%; chi-squared test, p=0.0001). The Phase 1/Phase 2 combination designation—which often signals uncertainty about whether a trial constitutes dose-finding or preliminary efficacy assessment—persists at 24% across all registration cohorts without temporal decline, suggesting that sponsors are not resolving early-stage ambiguity through sequential design refinement but rather perpetuating exploratory frameworks that defer the commitment to definitive efficacy testing. Registration volume has grown fourteen-fold between 2018 and 2024 (Figure 2A), yet this volumetric expansion has produced no proportional increase in Phase 2 or Phase 3 entries; the ratio of Phase 3 to Phase 1 trials (3:61, or 4.9%) compares unfavourably with the equivalent ratio in CAR-T (36:428, or 8.4%; Supplementary Figure 4A), indicating that the sEV field adds trials without advancing its pipeline toward the registration stages where regulatory decisions are made.

**Figure 2.**
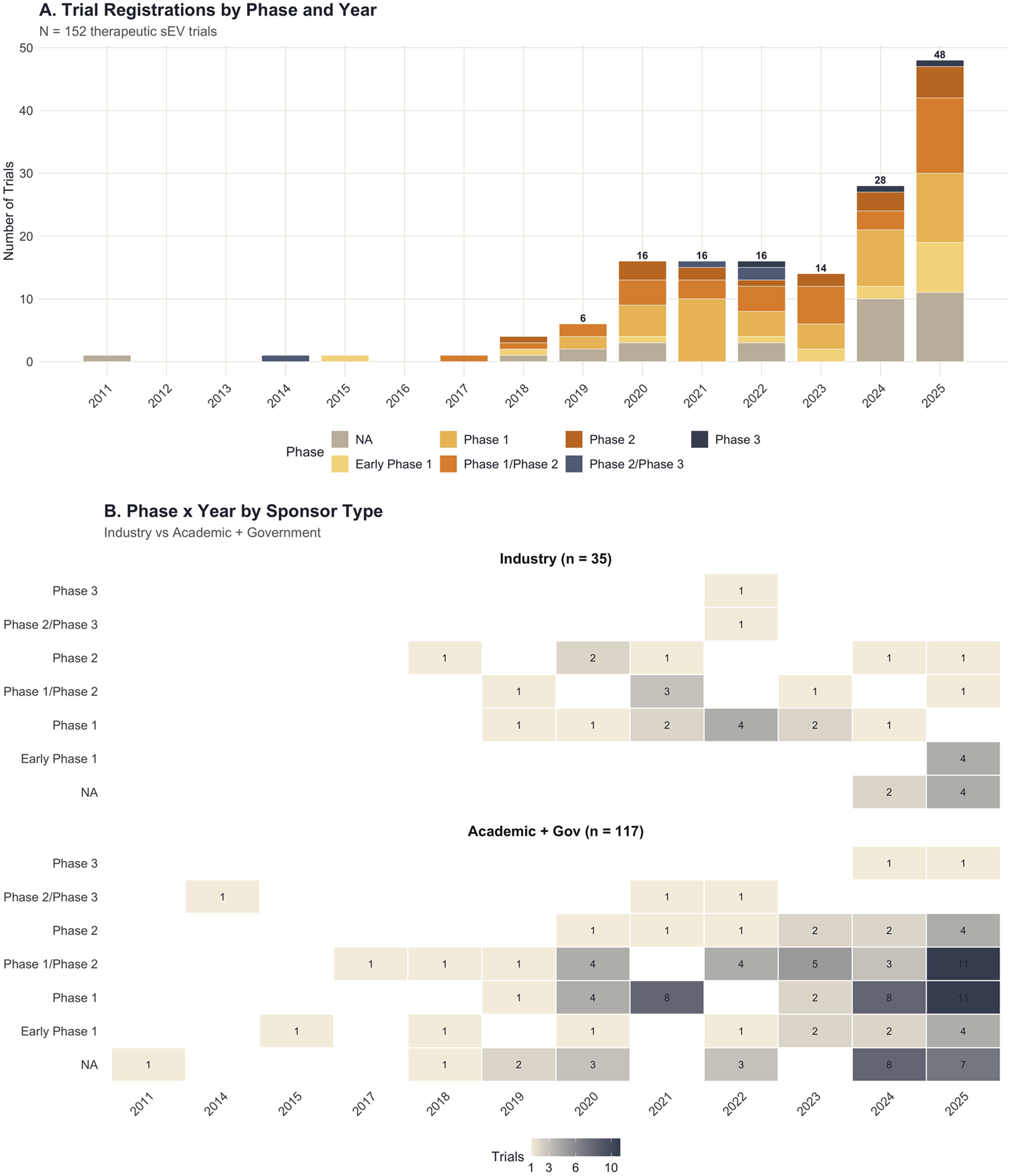
Phase Distribution of Therapeutic sEV Trials. (A) Stacked bar chart showing trial registrations by phase and year (2014–2025), illustrating fourteen-fold growth in volume without commensurate phase advancement. (B) Heatmap of trial counts by phase and year, stratified by sponsor type (Industry vs. Academic + Government), demonstrating that industry activity concentrates in early phases with minimal late-stage progression.

### The quality–failure paradox

To quantify methodological rigour across the therapeutic sEV pipeline, we constructed a composite quality index from four binary indicators assessable from registry records: data monitoring committee oversight, randomisation, blinding (single or double), and multi-arm design (Figure 3A). Industry-sponsored trials adopted each component at higher rates than academic programmes—DMC oversight (60% vs. 38%), randomisation (66% vs. 47%), blinding (57% vs. 37%), and multi-arm design (66% vs. 58%)—yielding a composite index of 2.49 versus 1.79 (Wilcoxon rank-sum p=0.016; ordinal regression OR=2.86, p=0.005), an approximately 39% elevation that was consistent across registration cohorts (Figure 3B).

**Figure 3.**
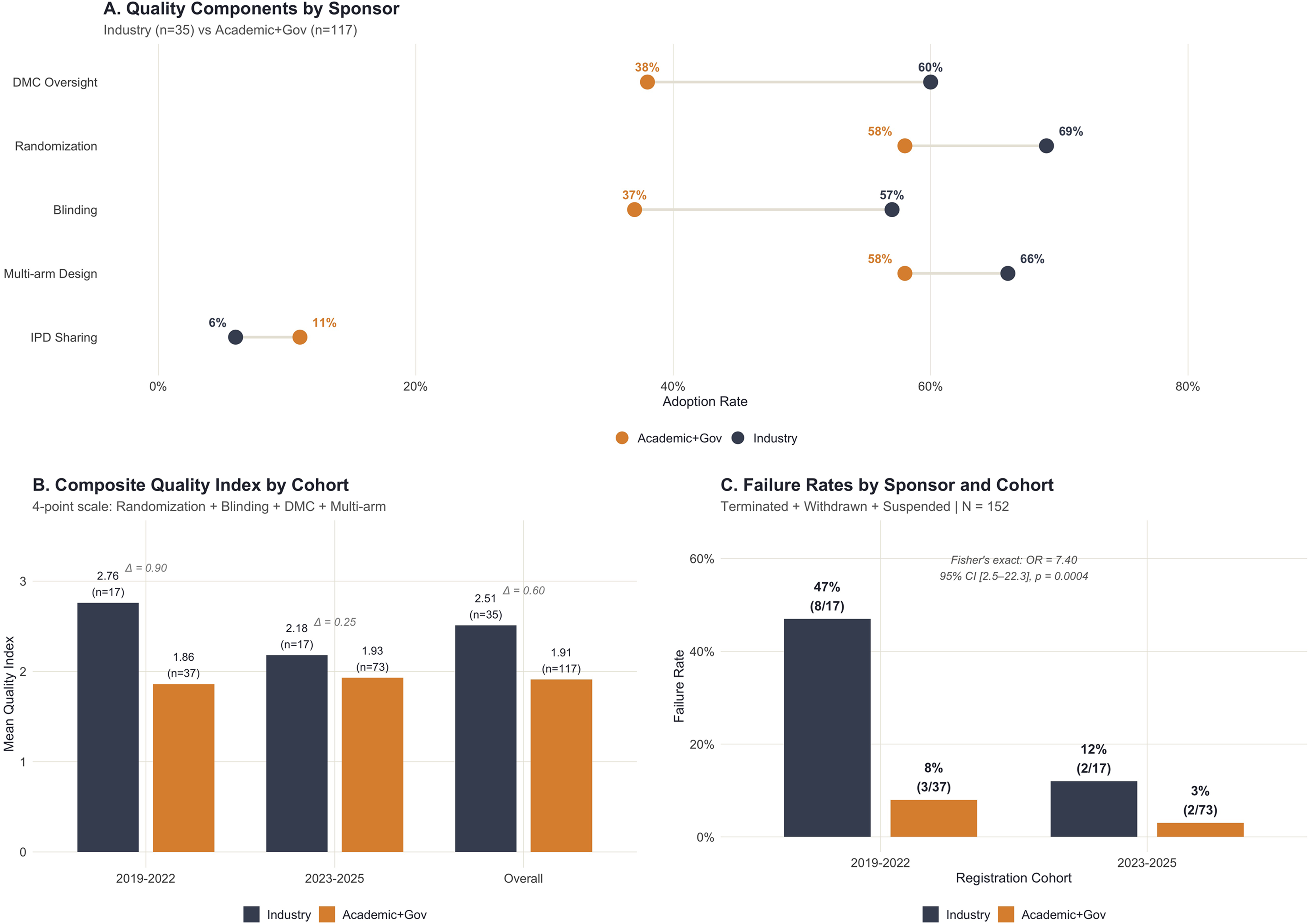
The Quality–Failure Paradox in Therapeutic sEV Trials. (A) Dumbbell plot of quality component adoption rates by sponsor type: DMC oversight (60% vs. 38%), randomisation (66% vs. 47%), blinding (57% vs. 37%), and multi-arm design (66% vs. 58%). (B) Composite quality index (4-point scale) by cohort and sponsor, with inter-group deltas annotated (Wilcoxon p=0.016; ordinal regression OR=2.86, p=0.005). (C) Failure rates by sponsor and registration cohort with Fisher’s exact test results (OR=7.40, 95% CI 2.5–22.3, p=0.0004).

This quality advantage, however, co-occurs with markedly higher failure rates. Industry-sponsored sEV trials have terminated, been withdrawn, or been suspended at a rate of 28.6% (10/35) compared with 5.1% (6/117) for academic programmes—a 5.6-fold differential evident across registration cohorts. Industry failure rates of 20% (1/5) in 2019–2020, 58% (7/12) in 2021–2022, and 12% (2/17) in 2023–2025, compared with academic rates of 18% (3/17), 0% (0/20), and 3% (2/73) in the same periods (Figure 3C). Fisher’s exact test yields an odds ratio of 7.40 (95% CI 2.5–22.3; p=0.0004), indicating that the association between industry sponsorship and failure substantially exceeds what chance alone would produce. Multivariable logistic regression adjusting for phase, year, and quality index confirms that the association persists after controlling for potential confounders (adjusted OR=11.64, p=0.0003), with quality index itself contributing no independent predictive value (OR=1.02, p=0.94).

Stratification by development stage revealed that the paradox concentrates in early clinical evaluation—industry-sponsored Phase 1 and Early Phase 1 trials failed at 40% (6 of 15) compared with 2.2% (1 of 46) for academic trials at the same stage (Fisher’s exact OR=30, p=0.0005), whereas the differential in later-phase trials did not reach significance (20% versus 7%, OR=3.3, p=0.10). This concentration is consistent with the interpretation that product definition gaps—most exposed during initial clinical evaluation, where characterisation deficiencies encounter regulatory and commercial scrutiny for the first time—drive industry termination decisions at the stage where the process-is-product constraint is first tested.

Three mechanisms—not mutually exclusive—explain this paradox. First, industry sponsors are more likely to implement formal go/no-go decision frameworks with pre-specified futility boundaries enforced by independent DMCs operating under charters that mandate interim analyses; these structures convert ambiguous efficacy signals into definitive stop decisions that register as formal failures in the trial registry, whereas academic programmes lacking such infrastructure may continue enrolling patients past the point at which futility would have been declared under a formal monitoring framework. Second, industry programmes disproportionately target demanding therapeutic indications where the existing standard of care sets a high bar for demonstration of incremental benefit, and where therapeutic effect sizes must exceed stringent evidentiary thresholds to justify programme continuation against opportunity cost. Third, the meta-epidemiological literature has long established that rigorous trial design—particularly adequate randomisation and blinding—yields smaller effect estimates for the same intervention (15), a phenomenon replicated across thousands of trials in Cochrane methodology reviews; quality infrastructure does not merely detect failure, it makes failure more likely for marginally effective interventions by removing the systematic biases (selection bias, performance bias, detection bias) that would otherwise inflate apparent efficacy in less rigorously designed evaluations.

A complementary finding supports this interpretation: among sEV trials that have formally failed, the mean quality index (2.31) exceeds that of completed trials (1.82), confirming that quality predicts failure detection rather than failure prevention. The implication is that the quality–failure paradox reflects an upstream problem—the premature entry of incompletely defined products into human testing—rather than a downstream methodological deficiency. Quality infrastructure functions as intended; it is the products entering these trials that are not yet ready to withstand rigorous evaluation.

### Sponsor composition and engagement dynamics

Industry engagement with sEV therapeutics remains significantly below CAR-T levels. Both modalities showed comparable early engagement (sEV 12%, CAR-T ex-China 18% in 2013–2016) and similar peak interest (sEV 38% in 2021–2022, CAR-T 42% in 2019–2020; p=0.70). However, by 2023–2025 the fields had diverged markedly (19% vs 33%; p=0.007). Unlike the more sustained CAR-T trajectory, sEV industry engagement proved non-monotonic, rising sharply before falling back to 19%— a pattern inconsistent with the prolonged industry commitment that typically precedes regulatory success in advanced therapies. The critical difference between sEV and CAR-T trajectories is not the shape—both modalities show post-peak decline—but the floor at which engagement stabilises: CAR-T maintains industry participation above 30%, sustained in part by revenue from approved products, whereas sEV industry share has fallen below 20% without an equivalent commercial anchor. This divergence reflects a key difference in trajectory: CAR-T’s first FDA approvals (2017–2018) provided a regulatory anchor that sustained industry engagement, whereas sEV therapeutics have yet to advance beyond early clinical stages, leaving industry without a comparable rationale to maintain commitment. In CAR-T, the first FDA approvals in 2017–2018 provided that anchor, whereas the sEV field’s inability to advance products beyond early clinical stages left industry engagement without a sustaining rationale.

The engagement gap becomes most consequential at late clinical stages. Of three sEV trials that have reached Phase 3, only one carries industry sponsorship—insufficient to constitute the kind of portfolio-level commitment required for registration. By contrast, 26 of 36 (72.2%) CAR-T Phase 3 trials in the ex-China population are industry-sponsored (Figure 4C). The academic-to-industry handoff that characterises successful translational trajectories—wherein academic proof-of-concept generates sufficient data to attract commercial development investment—has not occurred at scale in the sEV field; academic sponsors remain the dominant presence at every development stage, including those stages at which the capital requirements, regulatory expertise, and manufacturing scale of commercial sponsors become essential for registration success.

**Figure 4.**
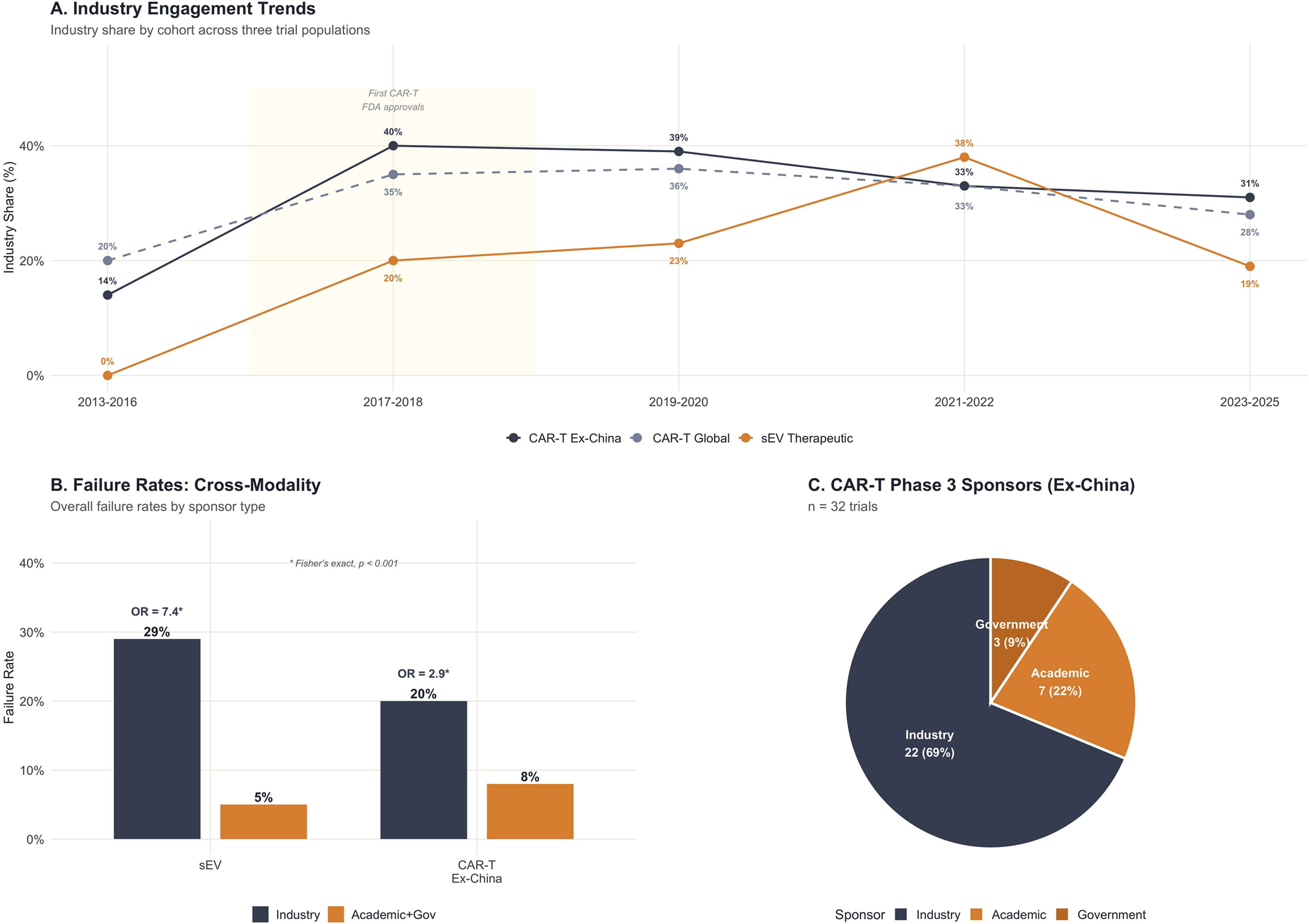
sEV vs CAR-T Developmental Trajectory. (A) Industry engagement trends across three trial populations (CAR-T Global, CAR-T Ex-China, sEV Therapeutic) by registration cohort; sEV industry share in the most recent cohort (19%) is significantly below CAR-T ex-China (33%, p=0.007). (B) Cross-modality failure rates by sponsor type with Fisher’s exact odds ratios annotated (sEV OR=7.4, CAR-T OR=2.5; both p<0.001). (C) Pie chart of CAR-T Phase 3 trial sponsors (ex-China), showing 69% industry dominance at registration stage.

### Cross-modality replication

To determine whether the quality–failure paradox is specific to sEV therapeutics or represents a broader structural phenomenon in complex biological therapies development, we analysed 1,131 CAR-T trials registered outside China (the ex-China restriction addresses the known heterogeneity in Chinese trial reporting, regulatory environment, and the distinct industry–academic dynamics of the Chinese biotechnology sector). Within the CAR-T ex-China population, industry-sponsored trials also fail at higher rates than academic programmes (20% vs. 9%), yielding a Fisher’s exact odds ratio of 2.51 (95% CI 1.8–3.6; p<0.001)—directionally concordant with the sEV result, though of smaller magnitude (Figure 4B). The replication of the industry–failure association with a modality that has achieved regulatory success is informative: it demonstrates that the paradox is not an artefact of sEV-specific failure but rather a general property of rigorous clinical development in complex biotherapeutic modalities, one that coexists with—rather than precludes—ultimate translational success when product definition is adequate.

Cochran–Mantel–Haenszel analysis pooling across modalities produces a common odds ratio of 2.76 (95% CI 1.64–4.67; p=1.8 × 10⁻), confirming that the industry–failure association is statistically robust after stratification.

Whether the paradox is equally strong in both modalities is less certain. The sEV odds ratio (7.4) is nearly threefold the CAR-T estimate (2.5), but a formal test for this difference — a sponsor × modality interaction term — did not reach significance (p = 0.067). Because a two-stratum comparison of this kind has limited statistical power, this result neither confirms that the paradox is stronger in sEV nor establishes that the two effects are equal; the apparent gap is suggestive rather than definitive. We therefore conclude that the direction of association replicates across modalities and that pooled analysis supports a significant common effect, but we cannot claim with confidence that the paradox operates at identical strength in both fields—the sEV odds ratio of 7.4 may genuinely exceed the CAR-T estimate of 2.5, potentially reflecting the more extreme product-definition deficit in the sEV pipeline where the majority of products lack the full mechanistic understanding of action and potency assay validation that even failed CAR-T programmes possessed at clinical entry.

### Registry abandonment and concentration risk

A substantial proportion of academic sEV therapeutic trials are listed as “Unknown” on ClinicalTrials.gov, reflecting registry abandonment (no updates within the mandated reporting window). Among the 152 therapeutic sEV trials, 24% of academic-sponsored programmes have Unknown status compared with only 9% of industry-sponsored programmes and 6% of CAR-T ex-China trials (p<0.0001).

This disparity directly affects interpretation of the quality–failure paradox. Reclassifying Unknown trials as failures raises the academic failure rate from 5.1% to ∼29%, collapsing the industry-versus-academic odds ratio from 7.40 to 1.40 and largely eliminating the paradox. Conversely, restricting the analysis to trials with confirmed outcomes preserves a robust association (OR=6.3). Consequently, the true non-productive rate among academic sEV programmes is approximately 29% (5% documented failures + 24% registry abandonment). Academic programmes thus fail silently— producing neither usable data nor definitive conclusions—whereas industry programmes generate interpretable termination signals that, while registering as formal failures, at least constrain the hypothesis space for subsequent development (16, 17).

Within the industry sector, trial outcomes are unevenly distributed: approximately one-quarter of all industry-sponsored sEV trials, and more than a half of industry failures originate from a small number of sponsors, and excluding the most heavily represented sponsor reduces the industry failure rate from 28.6% to 15.4%. This is still elevated relative to academic rates, but substantially less alarming than the headline figure suggests. Aggregate industry metrics are therefore disproportionately shaped by a small number of underperforming programmes rather than reflecting a uniform industry-wide pattern.

### Enrolment achievement

Among 46 therapeutic sEV trials for which paired planned and actual enrolment data could be extracted, overall enrolment achievement stood at 59% (1,091 of 1,859 planned participants; Figure 5). Academic trials substantially outperform industry in early-phase enrolment (93% vs. 40% in Phase 1), likely reflecting the smaller target sample sizes typical of academic investigator-initiated studies, and the relative ease of enrolling small safety cohorts at academic medical centres with established patient populations and pre-existing referral networks. Industry underperformance in enrolment is consistent with the more demanding inclusion criteria, multi-site coordination requirements, and competitive enrolment environments that characterise industry-sponsored protocols—features that simultaneously contribute to methodological quality and operational difficulty. The enrolment deficit is most pronounced in industry Phase 1/2 trials (3% achievement), which may reflect the premature termination of programmes before enrolment targets could be met rather than an inability to recruit *per se*; this interpretation aligns with the high industry failure rate and suggests that the enrolment metrics is a downstream consequence of product-level inadequacy too, rather than an independent operational barrier.

**Figure 5.**
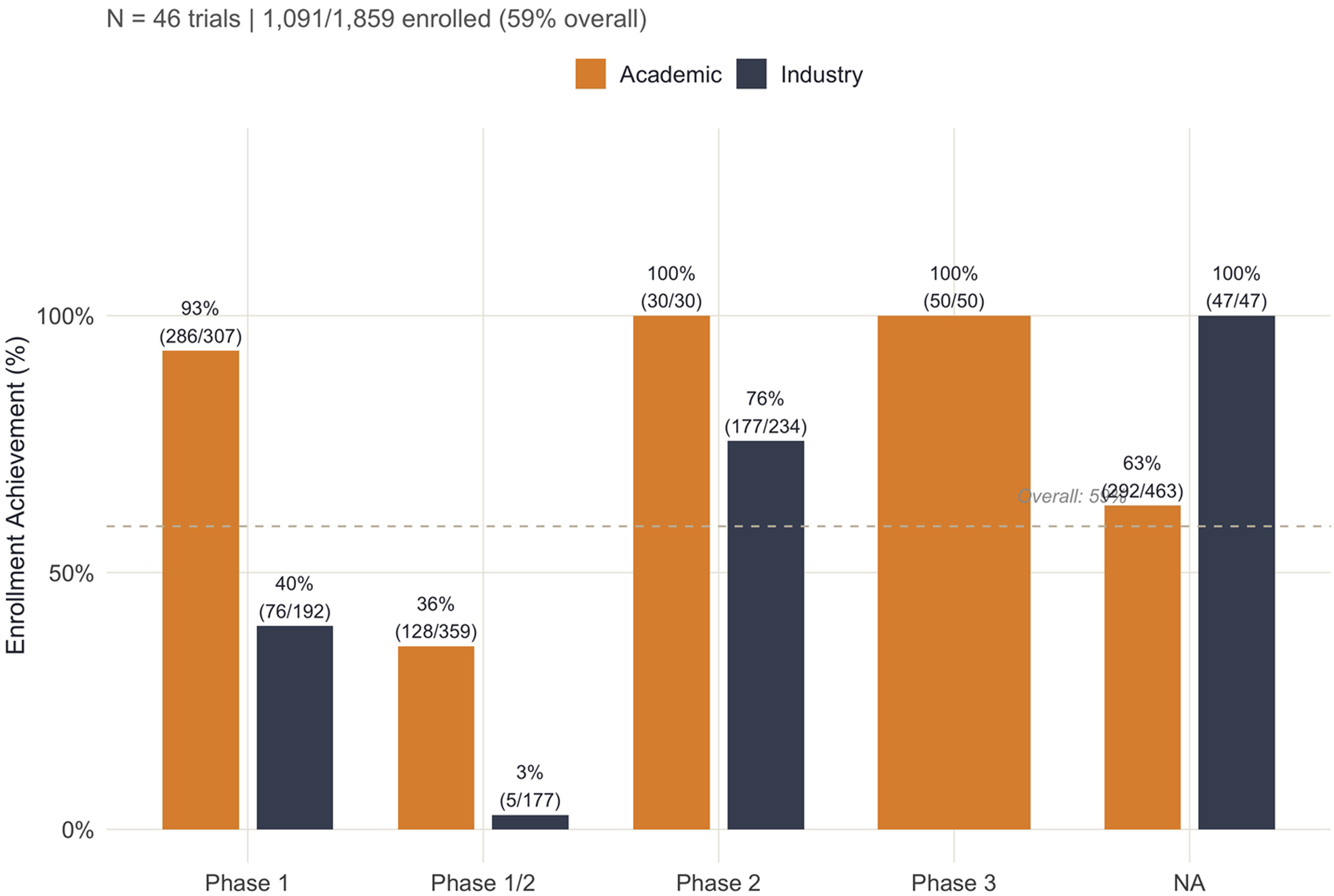
Enrolment Achievement by Phase and Sponsor. Bar chart showing enrolment achievement (actual/planned participants) across trial phases, stratified by sponsor type (N=46 trials with paired enrolment data). Overall achievement is 59% (1,091/1,859); academic programmes substantially outperform industry in Phase 1 enrolment (93% vs. 40%), reflecting smaller target sample sizes and single-centre convenience enrolment.

## DISCUSSION

The quality–failure paradox, as documented here, does not represent a dysfunction of methodological infrastructure but rather its proper functioning under conditions of premature clinical entry. Industry-sponsored sEV trials implement rigorous go/no-go frameworks, independent data monitoring, and pre-specified futility boundaries—and these structures perform exactly as designed, by identifying early that an incompletely defined product does not meet efficacy thresholds in a well-controlled evaluation. Typically, less controlled real-world settings would presumably make this default even more evident. The paradox persists only if one assumes that quality should prevent failure; once quality is understood as a detection mechanism—a lens through which inadequate products are identified and stopped before consuming further resources—the association between rigour and failure becomes not merely explicable but expected for any therapeutic platform where product definition lags behind clinical ambition.

The upstream constraint that our data collectively identify is a premature clinical entry without adequate product definition—a diagnosis reinforced by a registry abandonment analysis. When Unknown-status trials are incorporated into the failure calculus, the apparent concentration of failure in industry programmes dissolves; what remains is a field-wide non-productive rate of approximately 25–30% that manifests differently across sponsor types (formal termination in industry, silent abandonment in academia) but reflects the same underlying cause. Products entering clinical testing without locked manufacturing processes, validated potency assays, or characterised mechanisms of action will fail irrespective of trial design quality.

In this framing, registry abandonment represents not merely a reporting compliance failure, but a distinct mode of translational non-productivity with consequences that differ qualitatively from formal trial termination. A terminated trial, even one stopped for futility, generates data that constrains the hypothesis space and informs subsequent development decisions; an abandoned trial generates nothing. The 24% abandonment rate among academic sEV programmes represents a loss not only of the capital invested in those specific trials, but also of the potential information value that completed trials—whether positive or negative—would have contributed to collective understanding of sEV therapeutic potential. In a word - abandonment precludes the lessons learnt from well-conducted negative studies.

The ‘process is the product’ parallel with CAR-T therapy illuminates both the nature of this constraint and its resolution. CAR-T manufacturers resolved the product-identity challenge by treating each manufacturing process as generating a distinct product—despite both tisagenlecleucel and axicabtagene ciloleucel target CD19 with chimeric antigen receptors, they are two different products— and by locking processes sufficiently early in development to enable the lot-to-lot consistency that registration requires. The sEV field has not yet achieved this resolution (18-20). The majority of therapeutic trials employ EVs from mesenchymal stromal cells cultured under varying conditions, isolated by divergent methods (ultracentrifugation, size-exclusion chromatography, tangential flow filtration, polymer precipitation), and characterised by different analytical panels, making cross-trial comparison impossible and recognition of product distinctive protocols intractable. Until sEV developers adopt the product-specific development philosophy that enabled CAR-T regulatory success, the field will continue to generate volume without advancing toward registration. That the quality–failure paradox concentrates most acutely at the earliest clinical stage—where industry Phase 1 trials fail at 40% versus 2% for academic programmes—reinforces this diagnosis: it is at the point of first regulatory-grade evaluation that the absence of locked manufacturing processes and validated potency assays becomes operationally consequential, and it is at this stage that industry’s go/no-go discipline most efficiently identifies products that are not yet ready for human testing.

A parallel from lipid nanoparticle (LNP) development offers temporal perspective on platform maturation timelines that may temper expectations while clarifying the developmental path forward: liposomal formulations required approximately 30 years from initial clinical testing (Doxil approval, 1995) to achieve the manufacturing maturity and structure–function understanding that enabled rapid COVID-19 mRNA vaccine deployment in 2020–2021. The LNP platform succeeded not through volumetric trial expansion—early liposome trials were numerous but inconclusive—but through iterative process refinement, scale-up optimisation under GMP conditions, and the establishment of clear composition–activity relationships (lipid ionisability, PEG-lipid mole fraction, particle size distribution) that permitted rational design rather than empirical screening of formulation variants.

Fewer than 3% of all registered sEV clinical trials, making approximately 6% of therapeutic sEV trials, utilize engineered sEVs to deliver a specific therapeutic cargo as of late 2024 (5), leaving only a small fraction that can be directly compared to LNP formulations. Synthetic nanocarriers share with sEVs— biological nanocarriers—a diversity of carrier types (to a certain extent equivalent to sEV heterogeneity in terms of sources and composition), and a multitude of biomedical applications for which they have been historically proposed and tested. The two fields also share challenges that, regardless of their therapeutic benefit, slowed development and limited market appearance, including biological barriers, large-scale manufacturing complexity, biocompatibility and safety concerns, regulatory hurdles, and overall cost-effectiveness in comparison to established therapies. The lower and tunable compositional complexity of synthetic particles, and the absence of intrinsic biological activity, facilitated mechanistic understanding of their action and added value as a delivery modality. The sEV field, at roughly year 10 of clinical investigation is endowed by substantially less manufacturing standardisation than LNPs possessed at the same developmental stage, should not expect regulatory success from trial quantity alone; the path forward still requires investment in the kind of fundamental process understanding that preceded—rather than followed—LNP clinical success.

These findings carry implications for three stakeholder groups. For academic investigators, the data suggest that capital and patient resources allocated to early-phase trials of incompletely characterised products undermine returns; the 24% abandonment rate implies that nearly one in four academic sEV trials consumes resources without producing either a product or generalisable knowledge, representing an inefficient allocation of limited public research funding. Strategic investments should rather support the completion on convincing preclinical packages that would make subsequent clinical trials more likely to generate definitive answers. In line with this, for industry sponsors, the concentration-risk analysis demonstrates that the quality–failure paradox is partly shaped by idiosyncratic programme collapse within a small number of sponsors; the remaining industry portfolio fails at 15.4%, suggesting that product definition prior to clinical entry—rather than methodology deficit after entry—is the lever most likely to reduce attrition, and that investment in manufacturing process lock-down, potency assay validation, and mechanism-of-action characterisation before IND submission would yield higher returns than investment in trial size and rigor. For regulators and funding agencies, the persistent absence of phase designation in 20% of sEV trials and the 24% registry abandonment rate indicate inadequate oversight of trial registration compliance, and suggest that enforcement of existing reporting mandates, combined with clearer guidance on the product-characterisation expectations for EV-based therapeutics, would improve the field’s collective capacity to assess and advance its translational progress.

Several limitations constrain interpretation. Registry data provide an incomplete reflection of actual trial conduct—protocols may implement quality features not captured in registry fields, and conversely, registered features may not be faithfully implemented during trial execution. The composite quality index, while internally consistent and constructed from the most informative available indicators, cannot capture dimensions of methodological quality (biostatistical sophistication, endpoint selection appropriateness, manufacturing process control, analytical method validation) that may differ systematically between sponsor types, and can contribute substantively to trial success probability. The CAR-T comparator, although structurally motivated by the shared process-is-the-product constraint, involves a modality with a fundamentally different regulatory history and market dynamic; the existence of six approved products creates survivorship bias in interpretation of CAR-T failure rates, and the commercial success of early products generated revenue that funded subsequent pipeline expansion in ways not yet available to sEV developers. Geographic restriction of the CAR-T analysis to ex-China trials addresses known heterogeneity in reporting and regulatory environment but may introduce selection effects related to the distinct commercial landscapes of Western markets. Finally, the cross-sectional nature of trial registry data cannot distinguish between trials that will never complete and those that are merely delayed; some Unknown-status trials may ultimately report outcomes, though the duration of non-reporting (median exceeding 24 months for trials classified as Unknown in our dataset) suggests that the majority represent genuine abandonment rather than administrative delay.

## METHODS

### Data source and trial identification

Data were exported via the ClinicalTrials.gov API on December 18, 2025, using search terms “exosomes” (n=515) and “extracellular vesicles” (n=340). After removing 72 duplicates, 783 unique trials remained for analysis. The EU Clinical Trials Register was searched as a supplementary source, yielding only 23 additional sEV entries (predominantly diagnostic), confirming that ClinicalTrials.gov captures the majority of therapeutic programmes. For the CAR-T comparator, ClinicalTrials.gov was searched using “CAR-T,” yielding 2,337 interventional trials globally.

### Trial classification

EV trials were classified as therapeutic (interventional administration of EVs with efficacy or safety endpoints), diagnostic (EV-based biomarker or companion diagnostic), or other (cosmetic, nutritional, device-related). Classification was performed independently by both authors with disagreements resolved by consensus. Only therapeutic trials entered subsequent analyses. For the CAR-T population, we restricted to interventional trials administering CAR-T cells with therapeutic intent, excluding manufacturing-only or biomarker-only studies.

### Sponsor classification

Sponsors were classified as academic (university, hospital, research institute), industry (pharmaceutical, biotechnology, or medical device company), or government (national research agency, ministry of health). Collaborative trials were assigned to the lead sponsor as listed in the registry record. For analyses comparing industry versus non-industry, government-sponsored trials were pooled with academic programmes given their similar structural characteristics (investigator-initiated, publicly funded, non-commercial).

### Quality index construction

The composite quality index sums four binary indicators (range 0–4) assessable from trial registry records: (1) data monitoring committee or data safety monitoring board oversight, (2) randomisation (any type), (3) blinding (single-blind, double-blind, or triple-blind; open-label scored as 0), and (4) multi-arm design (two or more intervention arms, excluding single-arm studies). Each component was scored 1 if present and 0 if absent or not reported. This index prioritises trial-design features that meta-epidemiological research has identified as most strongly associated with effect estimate validity and that are consistently reportable from registry data across jurisdictions.

### Failure definition and capital efficiency rationale

Failure was defined as any trial registering a final status of Terminated, Withdrawn, or Suspended at the time of data extraction. This definition rests on a capital efficiency rationale: every trial that does not reach completion represents sunk investment (financial capital, patient exposure, investigator time) without the data return that would inform subsequent development decisions—regardless of the specific reason for non-completion. A trial that completes and misses its primary endpoint remains informative (it constrains the plausible effect-size space and may redirect subsequent development), whereas a terminated trial generates no such information. Unknown status (failure to update the registry within the mandated window) was treated as not-failed in the primary analysis; sensitivity analyses tested both reclassification of Unknown as failure and exclusion of Unknown-status trials from the denominator.

### Statistical analysis

Within each modality (sEV, CAR-T ex-China), the association between sponsor type (industry vs. academic/government) and trial failure was assessed using Fisher’s exact test for 2 × 2 contingency tables, with odds ratios and 95% confidence intervals reported. The composite quality index was compared between sponsor types using the Wilcoxon rank-sum test and ordinal logistic regression (proportional odds model). Multivariable logistic regression was performed to test whether the industry–failure association persisted after adjusting for phase, registration year, and quality index. The Cochran–Mantel–Haenszel (CMH) test was used to estimate a common odds ratio pooled across modalities; the Breslow–Day test assessed homogeneity of odds ratios across strata. Logistic regression with a sponsor × modality interaction term evaluated whether the paradox operates at different magnitudes across modalities. Sensitivity analyses included: (1) reclassification of all Unknown-status trials as failures, and (2) exclusion of Unknown-status trials from both numerator and denominator. All analyses were conducted in R (version 4.6.0). Statistical significance was set at α=0.05 (two-sided).

### CAR-T benchmark and geographic stratification

The CAR-T comparator population was stratified into global (all countries) and ex-China subsets to address known heterogeneity in Chinese trial reporting, regulatory environment, and industry– academic dynamics. The ex-China restriction removes approximately 45% of registered CAR-T trials, producing a population (n=1,131) more comparable to sEV therapeutics in regulatory environment and reporting standards.

## Data Availability

All data produced in the present study are available from public sources.

---

**Supplementary Figure 1.**
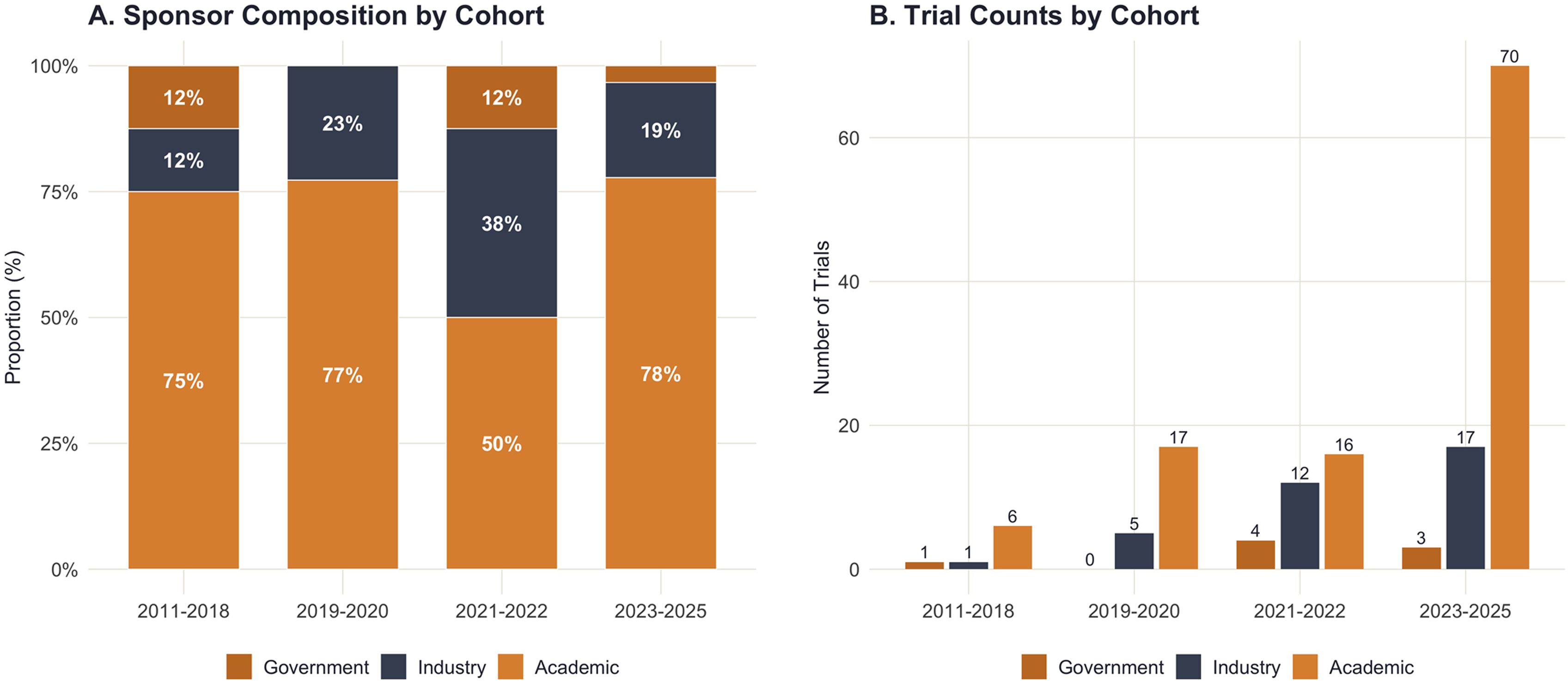
Temporal Sponsor Distribution of sEV Therapeutic Trials.

**Supplementary Figure 2.**
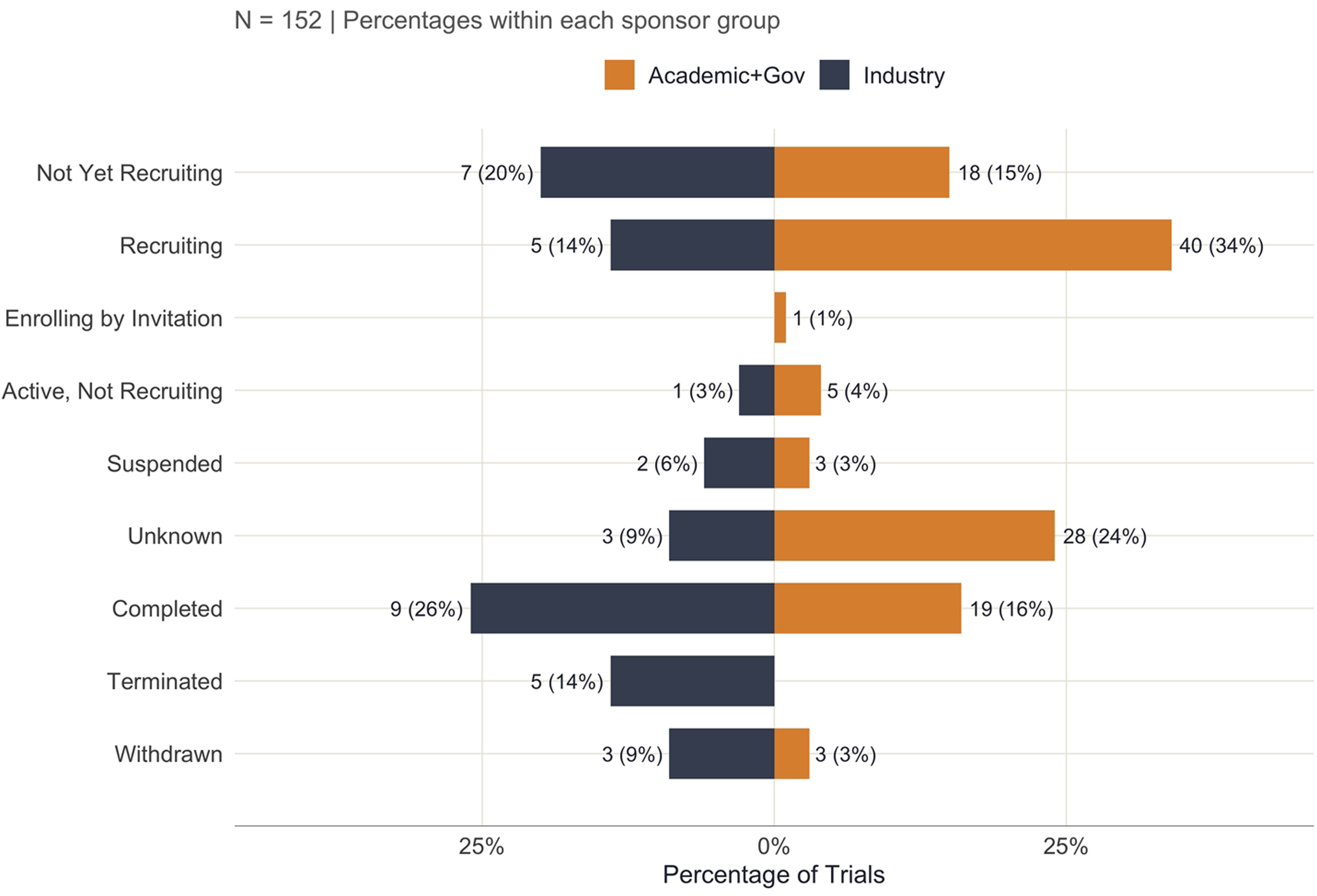
Trial Status by Sponsor Type.

**Supplementary Figure 3.**
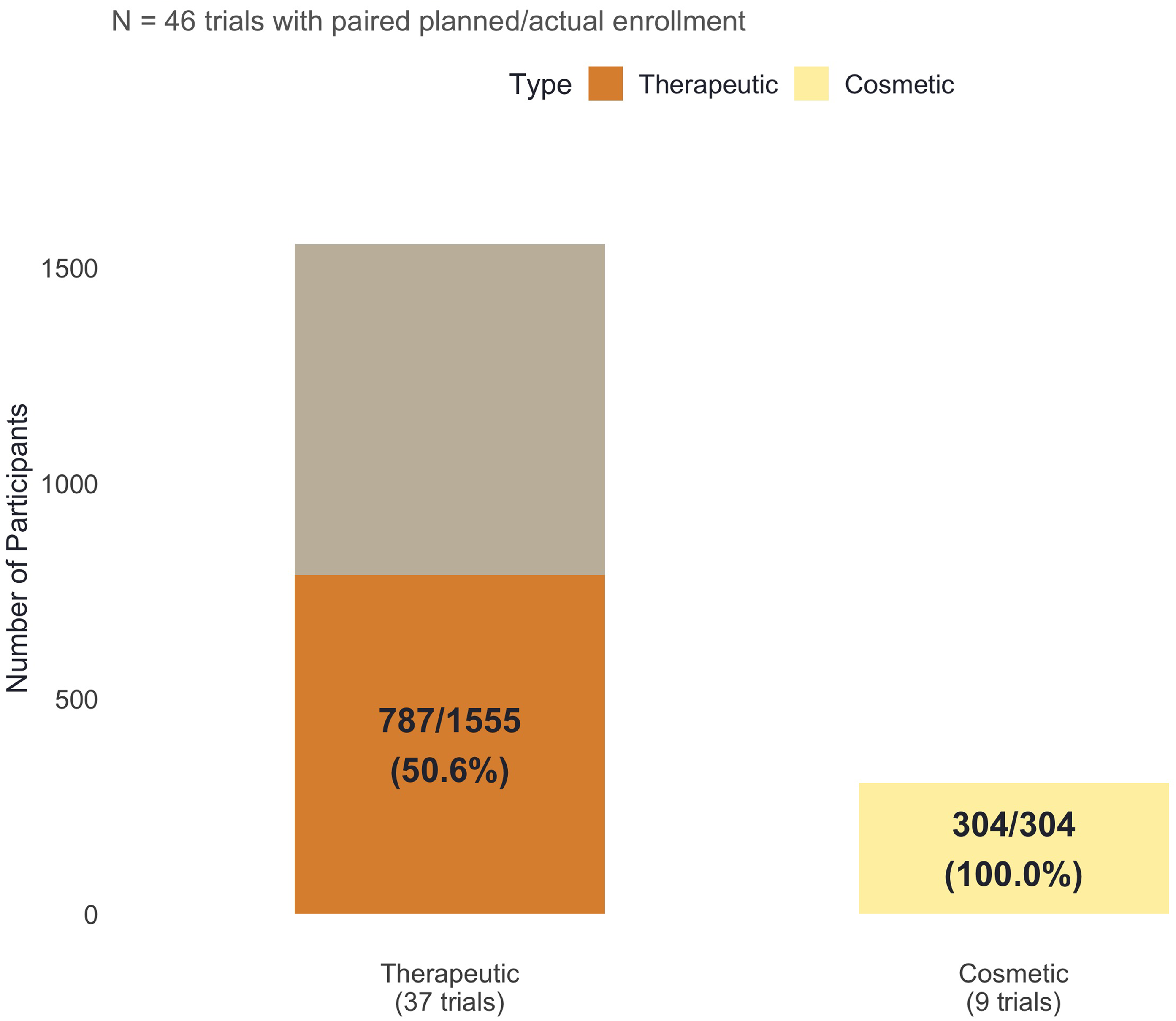
Enrollment: Therapeutic vs Cosmetic Trials.

**Supplementary Figure 4.**
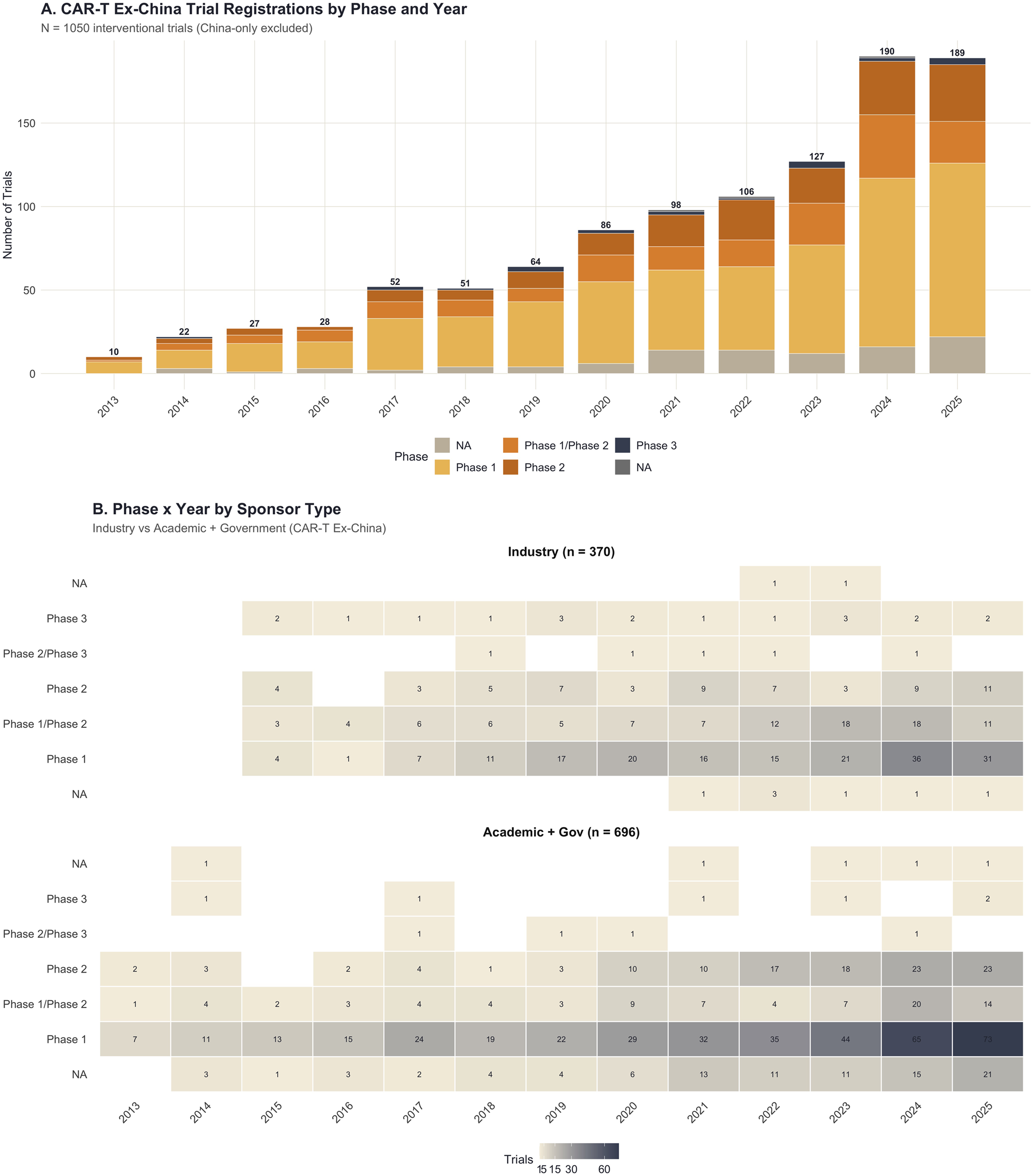
CAR-T Ex-China Phase Distribution.

## Notes

### Competing Interest Statement

The authors have declared no competing interest.

### Summary of Updates

reupload of figure files, initial version rendered figures too large in relationship to the text

